# Predicting the Clinical Management of Skin Lesions using Deep Learning

**DOI:** 10.1101/2020.11.02.20223941

**Authors:** Kumar Abhishek, Jeremy Kawahara, Ghassan Hamarneh

## Abstract

Automated machine learning approaches to skin lesion diagnosis from images are approaching dermatologist-level performance. However, current machine learning approaches that suggest management decisions rely on predicting the underlying skin condition to infer a management decision without considering the variability of management decisions that may exist within a single condition. We present the first work to explore image-based prediction of clinical management decisions directly without explicitly predicting the diagnosis. In particular, we use clinical and dermoscopic images of skin lesions along with patient metadata from the Interactive Atlas of Dermoscopy dataset (1,011 cases; 20 disease labels; 3 management decisions) and demonstrate that predicting management labels directly is more accurate than predicting the diagnosis and then inferring the management decision (13.73 ± 3.93% and 6.59 ± 2.86% improvement in overall accuracy and AUROC respectively), statistically significant at *p <* 0.001. Directly predicting management decisions also considerably reduces the over-excision rate as compared to management decisions inferred from diagnosis predictions (24.56% fewer cases wrongly predicted to be excised). Furthermore, we show that training a model to also simultaneously predict the seven-point criteria and the diagnosis of skin lesions yields an even higher accuracy (improvements of 4.68 ± 1.89% and 2.24 ± 2.04% in overall accuracy and AUROC respectively) of management predictions. Finally, we demonstrate our model’s generalizability by evaluating on the publicly available MClass-D dataset and show that our model agrees with the clinical management recommendations of 157 dermatologists as much as they agree amongst each other.

## Introduction

Until a few years ago, the computer-aided diagnosis of skin lesions from images involved extracting the lesion boundary to distinguish it from the surrounding healthy skin (i.e., skin lesion segmentation), followed by calculating features based on rules developed by dermatologists such as the ABCD rule and the CASH rule^1,2^ based on the obtained segmentation, and ultimately using these features to train classical machine learning models (e.g., support vector machines and random decision forests^3–8^) to recommend diagnoses. Since skin lesion segmentation is an intermediate task in the dermatological analysis pipeline, the use of deep learning to predict diagnosis directly from the images, bypassing the segmentation, is now commonplace^9–12^ and is evident in other imaging modalities as well^13–17^. We project a similar trend where the model deemphasizes predicting the diagnosis and instead prioritizes producing accurate predictions of the ultimate clinical task (e.g., clinical management).

While deep learning based diagnoses of dermatological conditions from images are reaching the performance levels of medical professionals^9,10,18,19^, no work has been published to directly predict the management of the disease. Even in scenarios where the diagnosis is decided by an automated prediction model, the general physician or the dermatologist must still decide on the disease management (be it the treatment plan or some other course of action, e.g., requesting other exams or future follow-ups). Moreover, in some cases, accurately diagnosing the underlying skin condition may not be possible from an image alone. For example, a recent study evaluating the ‘majority decision’ obtained from over a hundred dermatologists for melanoma classification resulted in a sensitivity of 71.8% with respect to the ground truth diagnosis^20^. Thus, in the case where the visual presentation of a lesion is ambiguous, rather than diagnosing the condition, the correct action may be to perform a biopsy to gain further information. Machine learning-based approaches that classify the underlying skin condition and use the predicted skin condition to directly decide on a disease management (e.g., Han et al.^21^) may not well distinguish among different management decisions that exist within a single class. A management decision (e.g., scheduling a follow-up visit to monitor the skin lesion progression) may even be necessary to confirm a diagnosis (when there is insufficient information within the image), and therefore must precede it. For example, the clinical management decision for a nevi without atypical characteristics may be that no further action is required, whereas for a nevi with atypical characteristics, a dermatologist may opt for a clinical follow up or an excision, which may depend on the severity of the atypical characteristics. Therefore, it is desirable to explore the performance of an artificial intelligence based automatic skin disease management prediction system. Such a system can suggest management decisions to a clinician (i.e., as a second opinion) or directly to patients in under-served communities^22^. Moreover, when there are fewer management decisions to choose from than there are diagnosis classes (since multiple subsets of disease classes may be prescribed the same course of action), predicting the management decisions is likely a simpler computational problem to address than predicting the diagnosis and then inferring the management.

Previous work on clinical management prediction for skin lesions includes comparing management predictions made by MelaFind (a handheld imaging device developed by MELA Sciences Inc. which acquires 10 spectral bands) to histologic slides as the reference labels^23^ and to decisions made by dermatologists^23,24^. Carrara et al.^25^ used shallow artificial neural networks to predict whether a skin lesion should be excised based on lesion descriptors extracted from their multispectral images (15 spectral bands). Marchetti et al.^26^ compared the diagnostic accuracy of an ensemble of automated diagnosis prediction methods (including 2 machine learning-based methods) to the management decisions made by 8 dermatologists for a set of 100 dermoscopic images, but did not directly predict the management decisions using a learning-based approach. To the best of our knowledge, we are the first to predict management decisions, without relying on explicit diagnosis predictions, using machine learning (shallow or deep) using only RGB images of skin lesions and, in fact, using a deep learning-based approach for any skin lesion imaging modality. We evaluate our proposed method on the Interactive Atlas of Dermoscopy Dataset^27–29^, the largest publicly available database containing both dermoscopy and clinical skin lesion images with the associated management decisions, and show that predicting management decisions directly is more accurate than inferring the management decision from a predicted diagnosis. We also validate our model on the publicly available Melanoma Classification Benchmark (MClass-D)^18,30^ and show that our model exhibits excellent generalization performance when evaluated on data from a different source, and that our model’s clinical management predictions are in agreement with those made by 157 dermatologists.

## Results and Discussion

The Interactive Atlas of Dermoscopy dataset was used to test the performance of a model trained to predict the clinical management decisions (MGMT_pred_) compared with inferring the management decisions based on the outputs of a diagnosis prediction model (MGMT_infr_). This dataset contains 1,011 lesion cases spanning 20 diagnosis labels (Table 1) grouped into 5 categories^28^: basal cell carcinoma (BCC), nevus (NEV), melanoma (MEL), seborrheic keratosis (SK), and others (MISC), and 3 management decisions: ‘clinical follow up’ (CLNC), ‘excision’ (EXC), and ‘no further examination’ (NONE). The MClass-D dataset^30^ was used to compare the diagnosis and the management prediction performance of our model with that of dermatologists. This dataset contains 100 dermoscopic images comprising of 80 benign nevi and 20 melanomas, as well as the responses of 157 dermatologists when asked to make a clinical management decision to each of these 100 images: ‘biopsy/further treatment’ (EXC) or ‘reassure the patient’ (NOEXC).

**Table 1.** Breakdown of the seven-point criteria evaluation dataset^29^ by management and diagnosis labels and the train-valid-test splits used to train the model.

| Management Labels | Diagnosis Labels |  |  |  |  | Split |  |  | Total |
| --- | --- | --- | --- | --- | --- | --- | --- | --- | --- |
|  | BCC | NEV | MEL | MISC | SK | Training | Validation | Testing |  |
| CLNC | 0 | 268 | 0 | 24 | 4 | 133 | 51 | 112 | 296 |
| EXC | 42 | 278 | 252 | 23 | 10 | 235 | 127 | 243 | 605 |
| NONE | 0 | 29 | 0 | 50 | 31 | 45 | 25 | 40 | 110 |
| **Total** | 42 | 575 | 252 | 97 | 45 | 413 | 203 | 395 |  |

### Interactive Atlas of Dermoscopy Dataset

#### Predicting whether a lesion should be excised or not

The outputs of the diagnosis prediction model are mapped to a binary management decision (MGMT_infr, binary_; Figure 1 (a1)) of whether a lesion should be excised (EXC) or not (NOEXC). All malignancies (MEL and BCC) are mapped to EXC and all other diagnoses to NOEXC. Similarly, the outputs of the management prediction model are mapped to a binary decision (MGMT_pred, binary_; Figure 1 (b1)) by retaining the EXC class from MGMT_pred_ as is and grouping CLNC and NONE to form NOEXC. These binary mapping-based approaches serve as our baselines, and we observe that MGMT_infr, binary_ correctly predicts 218 of the 395 cases (overall accuracy = 55.19%), whereas MGMT_pred, binary_ yields a superior classification performance of 289 correct predictions (overall accuracy = 73.16%), outperforming the inference-based management decision by 17.97%.

**Figure 1.**
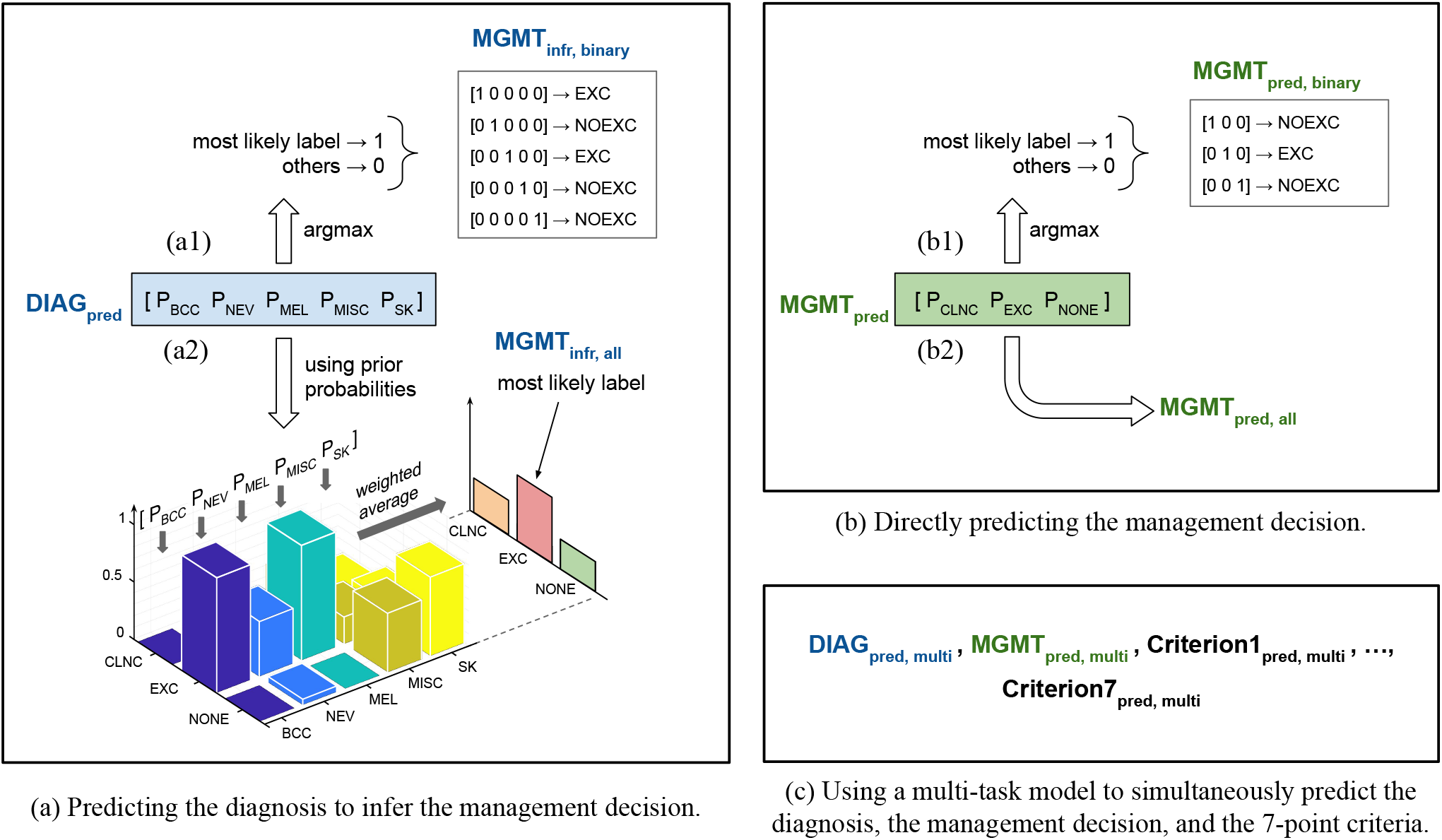
An overview of the three prediction models. All the models take the clinical and the dermoscopic images of the skin lesion and the patient metadata as input. Note that we also perform an input ablation study (Section “A multi-task prediction model”; Table 4). (a) The first model predicts the lesion diagnosis probabilities, DIAG_pred_. (b) The second model predicts the management decision probabilities, MGMT_pred_. (c) The third is a multi-task model and predicts the seven-point criteria (Criterion{1, 2, …, 7}_pred, multi_) in addition to DIAG_pred, multi_ and MGMT_pred, multi_. The argmax operation assigns 1 to the most likely label and 0 to all others. For (a), DIAG_pred_ diagnosis is used to arrive at a management decision either using (a1) binary labeling, MGMT_infr, binary_, or (a2) prior based inference, MGMT_infr, all_. Similarly, the outputs of (b) can be used to directly predict a management decision using either (b1) binary labeling, MGMT_pred, binary_, or (b2) all the labels, MGMT_pred, all_. As explained in the text, the diagnosis labels are basal cell carcinoma (BCC), nevus (NEV), melanoma (MEL), seborrheic keratosis (SK), and others (MISC), and the management decision labels are ‘clinical follow up’ (CLNC), ‘excision’ (EXC), and ‘no further examination’ (NONE). In the case of binary management decisions, we predict whether a lesion should be excised (EXC) or not (NOEXC).

#### A data-driven approach to inferring management decision from diagnosis predictions

Since cases belonging to a disease label can be managed in multiple ways, a data-driven approach using conditional probabilities (Equation (3)) can be adopted to infer the probabilistic management decisions from the diagnosis predictions, and this does not have to be restricted to a binary management. These inferred management decisions (MGMT_infr, all_; Figure 1 (a2)) can then be compared to the probabilistic outputs of the management prediction model (MGMT_pred, all_; Figure 1 (b2)).

Figure 2(a) shows the distribution of the four sets of distance measures for examining the correctness of the probabilistic MGMT_infr, all_ and MGMT_pred, all_ predictions with respect to the target labels, where each dot represents a test case. For (1 - cosine similarity), the mean [95% CI] distance is lower for MGMT_pred, all_ as compared to MGMT_infr, all_ (0.3584 [0.3260 - 0.3909] versus 0.4703 [0.4490 - 0.4915]; Cohen’s d = 0.4033). We observe similar patterns for the Jensen-Shannon divergence (0.3551 [0.3320 - 0.3783] versus 0.4397 [0.4249 - 0.4544]; Cohen’s d = 0.4311), the Wasserstein distance (0.1358 [0.1246 - 0.1469] versus 0.2687 [0.2581 - 0.2793]; Cohen’s d = 1.2064), and the Hellinger distance (0.4131 [0.3868 - 0.4394] versus 0.5111 [0.4944 - 0.5278]; Cohen’s d = 0.4404).

**Figure 2.**
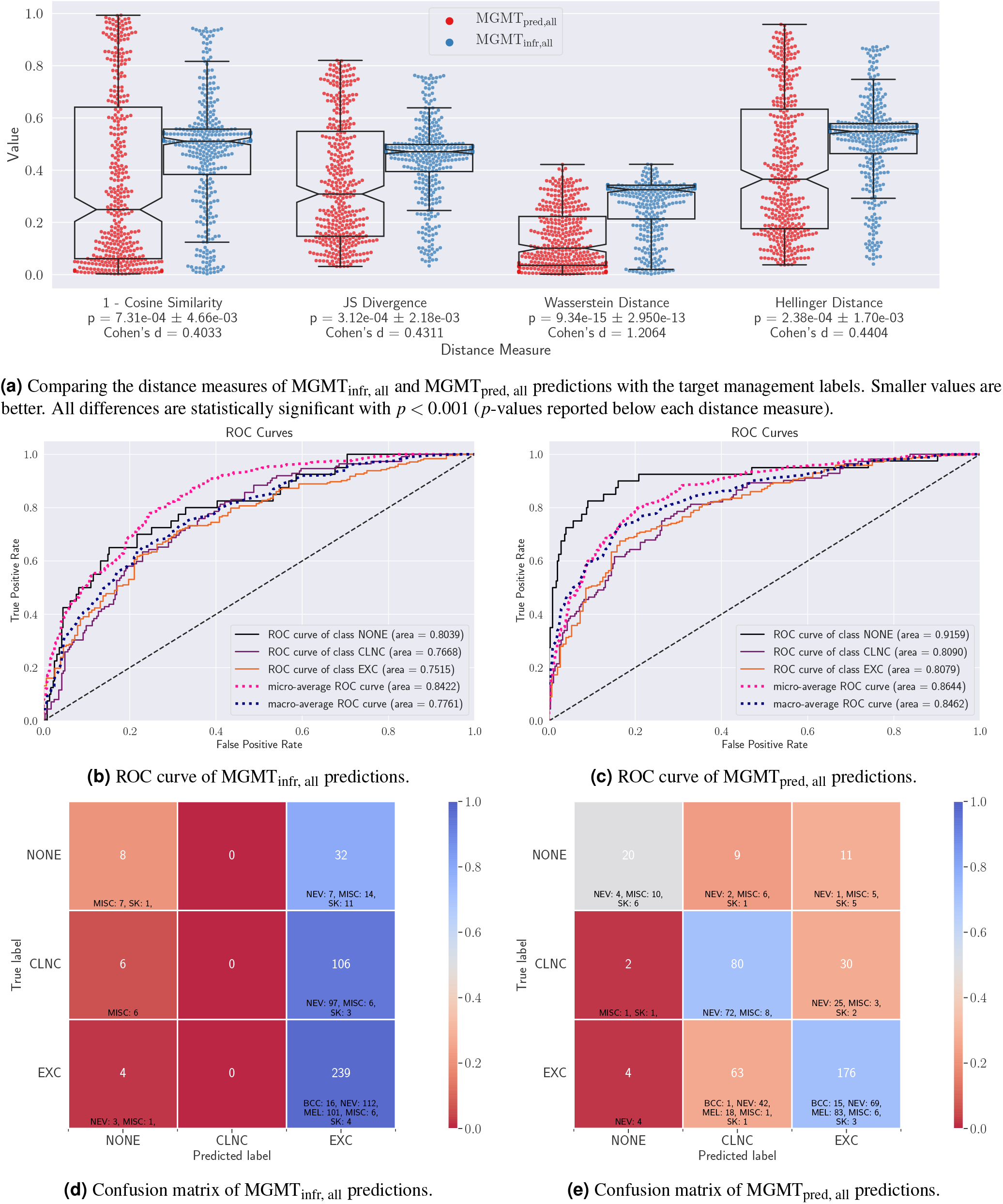
Quantitative evaluation of the MGMT_infr, all_ and MGMT_pred, all_ predictions. (a) Violin plots of the distance measures of the probabilistic predictions show that the MGMT_pred, all_ predictions are closer (statistically significant) to the target labels for test data. (b, c) ROC curves and (d, e) confusion matrices of MGMT_infr, all_ and MGMT_pred, all_ respectively along with cell-wise diagnosis breakdown. Note that MGMT_infr, all_ has a tendency to over-excise lesions.

The final management predictions from the two approaches (MGMT_infr, all_ and MGMT_pred, all_) are obtained by extracting the most likely label over the probabilistic predictions, and their quantitative results are presented in Table 2. The ROC curves for the two approaches are shown in Figure 2 (b, c) and their respective confusion matrices, with each cell in the confusion matrices also indicating a diagnosis-wise breakdown of the test samples, are shown in Figure 2 (d, e).

**Table 2.** Comparing skin lesion management prediction results obtained using MGMT_infr, all_ and MGMT_pred, all_. All the prediction models have been trained using all the input data modalities (i.e., clinical image, dermoscopic image, and patient metadata). Mean ± standard deviation reported for all the metrics for the 3-fold cross validation.

| Management Labels | MGMT<sub>infr, all</sub> |  |  |  |  | MGMT<sub>pred, all</sub> |  |  |  |  |
| --- | --- | --- | --- | --- | --- | --- | --- | --- | --- | --- |
|  | Sensitivity | Specificity | Precision | AUROC | Overall Accuracy | Sensitivity | Specificity | Precision | AUROC | Overall Accuracy |
| NONE | 0.2 | 0.9718 | 0.4444 | 0.8039 | - | 0.5 | 0.9831 | 0.7692 | 0.9159 | - |
| CLNC | 0.0 | 1.0 | 0.0 | 0.7668 | - | 0.7143 | 0.7456 | 0.5263 | 0.8090 | - |
| EXC | 0.9835 | 0.0921 | 0.634 | 0.7515 | - | 0.7243 | 0.7303 | 0.8111 | 0.8079 | - |
| **Average** | 0.3945 | 0.6880 | 0.3595 | 0.7741 | 0.6253 | **0.6462** | **0.8196** | **0.7022** | **0.8443** | **0.6987** |
| **3-Fold Cross Validation** | 0.3943<br>± 0.0025 | 0.6871<br>± 0.0018 | 0.3829<br>± 0.0172 | 0.7758<br>± 0.0178 | 0.6139<br>± 0.0198 | **0.6033**<br>± **0.0369** | **0.8107**<br>± **0.0093** | **0.7104**<br>± **0.0184** | **0.8266**<br>± **0.0126** | **0.6974**<br>± **0.0018** |

We observe that the overall accuracy and AUROC of MGMT_infr, all_ (62.53% and 0.7741) are considerably lower than those of MGMT_pred, all_ (69.87% and 0.8443), indicating that predicting the management decisions directly leads to a better accuracy than predicting the diagnosis and then inferring the management. Another interesting observation is that MGMT_infr, all_ predictions tend to favor EXC (excision) more than other labels (as can be observed by the dominant blue colored cells in the rightmost column of Figure 2(b)), which although leads to an excellent sensitivity (0.9835) for the EXC class, yields unacceptable classification performance for the other two classes (0.2 and 0.0, for NONE and CLNC respectively). For example, none of the clinical follow-up cases were predicted correctly by MGMT_infr, all_, and 106 cases (94.64%) were predicted to be over-treated by excision. Similarly, the algorithm wrongly predicted excising 32 cases (40%) that, in fact, needed no further examination. On the other hand, MGMT_pred, all_ yields a higher overall accuracy without favoring any particular class. Finally, three-fold cross validation results show that this improvement in performance holds true for all metrics across multiple training, validation, and testing partitions of the dataset, with sufficiently low standard deviations across all folds.

#### A multi-task prediction model

It has been shown that models optimized to jointly predict related tasks perform better than models trained on individual tasks separately^31^. As such, we expect to observe an improvement in the management prediction accuracy of our multi-task model trained to simultaneously predict the seven-point criteria^32^ of the lesions (Criteria1_pred, multi_ … Criteria7_pred, multi_), the diagnosis label (DIAG_pred, multi_), and the management decision (MGMT_pred, multi_). We plot the confusion matrix and the ROC curves for MGMT_pred, multi_ for this multi-task model in Figure 3(a). As expected, we improve the overall management prediction accuracy by 3.8% (from 69.87% to 73.67%). Moreover, since we have fairly imbalanced classes (see Table 1; for example, there are 243 EXC cases as compared to only 40 NONE cases in the test partition) where ROC curves can indicate an “overly optimistic view” of the algorithm’s performance^33^, we also plot the precision-recall curves for the multi-task model in Figure 3(b). A detailed analysis of class-wise performance is presented in Table 3. Three-fold cross validation results show the robustness of this multi-task model to different training, validation, and testing partitions of the dataset. In addition to its higher management prediction accuracy, this multi-task model may be regarded as less opaque and more trustworthy as its final management prediction was linked to clinically meaningful predictions, i.e., the seven-point criteria and the diagnosis. Finally, an input data ablation study for estimating the importance of each input modality (i.e., clinical image, dermoscopic image, and patient metadata) was conducted where six prediction models were trained using various combinations of input data modalities. Their quantitative results presented in Table 4 and the *p*-values for pairwise comparison of their predictions using mid-*p* McNemar’s test are as shown in Figure 4. We draw the following key observations:

**Table 3.**
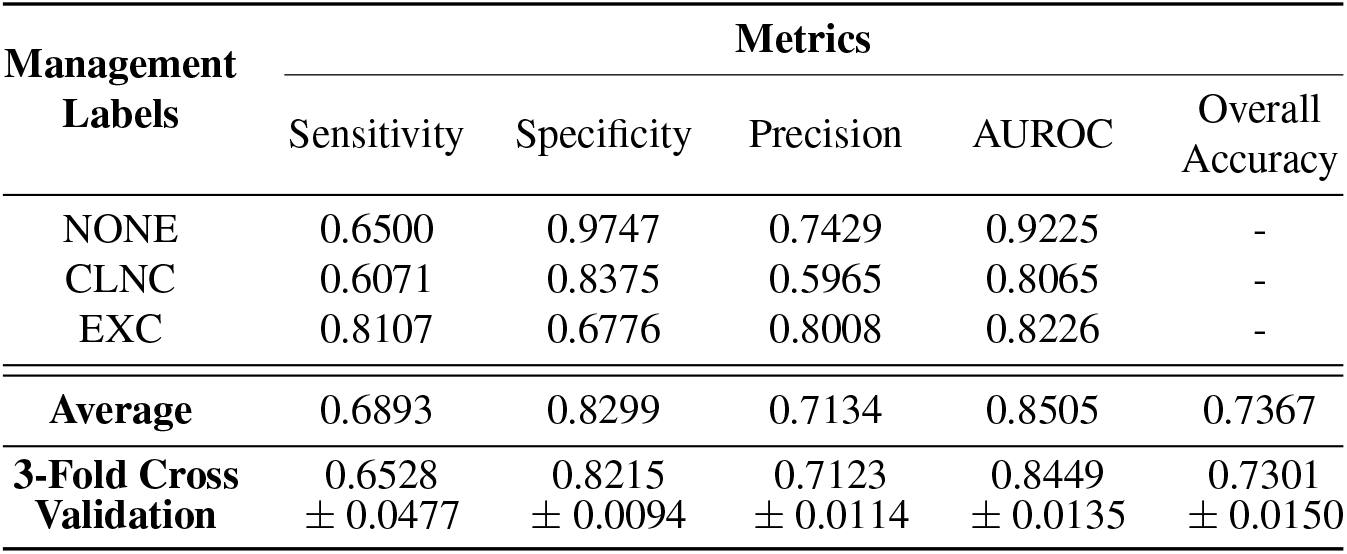
Skin lesion management prediction results MGMT_pred, multi_ obtained using a multi-modal multi-task model. All the prediction models have been trained using all the input data modalities (i.e., clinical image, dermoscopic image, and patient metadata). Mean ± standard deviation reported for all the metrics for the 3-fold cross validation.

**Table 4.** Input data modality ablation study for skin lesion management prediction results MGMT_pred, multi_ obtained using a multi-task model. Each experiment is named so as to denote the input data modalities it uses to make the management predictions, and ‘C’, ‘D’, and ‘M’ refer to clinical image, dermoscopic image, and patient metadata, respectively.

| Experiment Name | Input Data |  |  | Metrics |  |  |  |  |
| --- | --- | --- | --- | --- | --- | --- | --- | --- |
|  | Clinical Image | Dermoscopic Image | Patient Metadata | Sensitivity | Specificity | Precision | AUROC | Overall Accuracy |
| C | ✓ | ✗ | ✗ | 0.5997 | 0.7911 | 0.5466 | 0.7781 | 0.6051 |
| CM | ✓ | ✗ | ✓ | 0.6050 | 0.7983 | 0.5684 | 0.7852 | 0.6405 |
| D | ✗ | ✓ | ✗ | 0.6935 | 0.8384 | 0.6265 | 0.8630 | 0.6962 |
| DM | ✗ | ✓ | ✓ | **0.7126** | **0.8424** | 0.6622 | **0.8644** | 0.7215 |
| CD | ✓ | ✓ | ✗ | 0.5830 | 0.8060 | **0.7393** | 0.8335 | 0.7342 |
| CDM | ✓ | ✓ | ✓ | 0.6893 | 0.8299 | 0.7134 | 0.8505 | **0.7367** |

**Figure 3.**
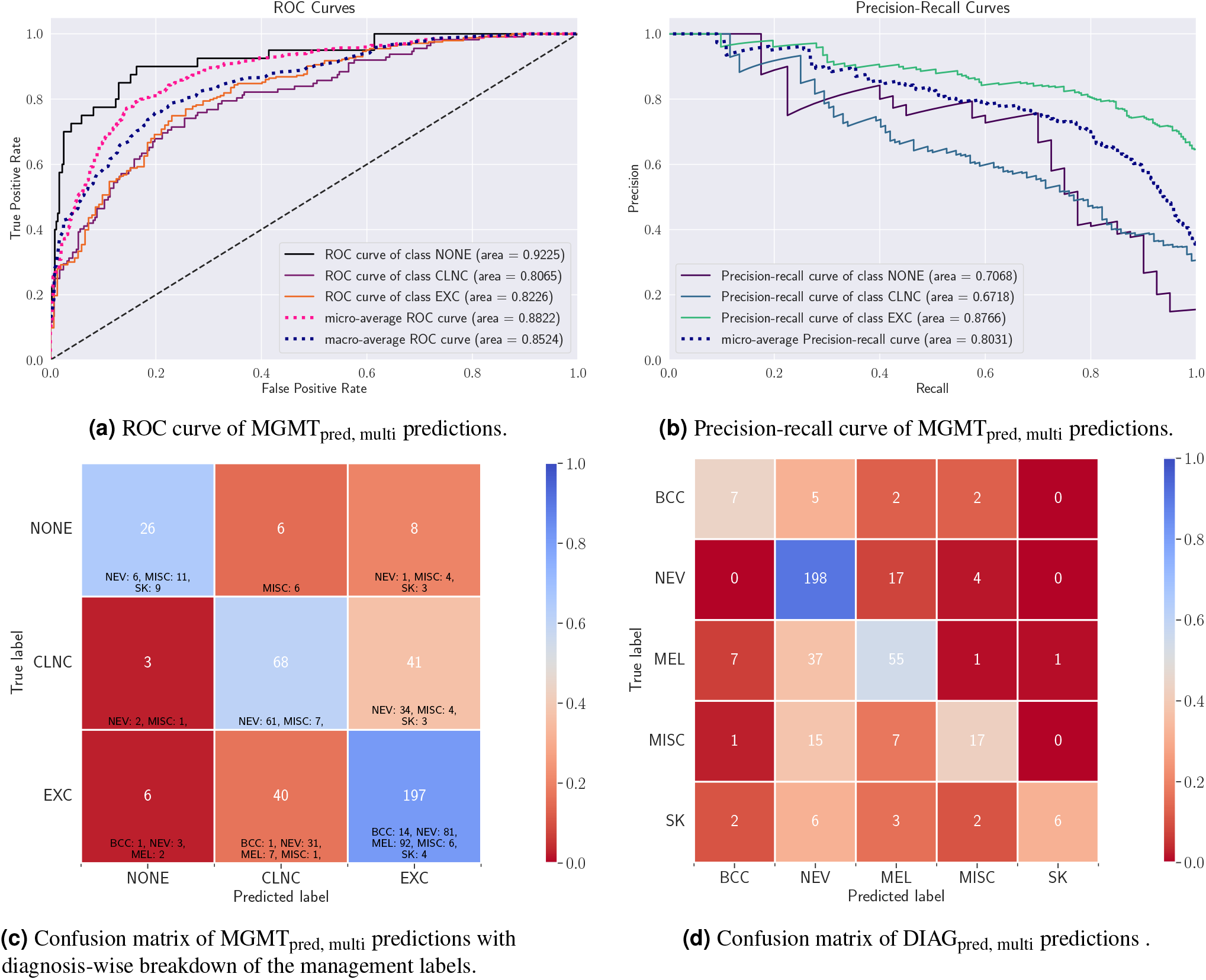
Evaluating the multi-modal multi-task model. (a) ROC curve and (b) precision-recall curve for the management prediction task. Confusion matrices for (c) the management prediction task and (d) the diagnosis prediction task along with the diagnosis-wise breakdown for the management labels.

**Figure 4.**
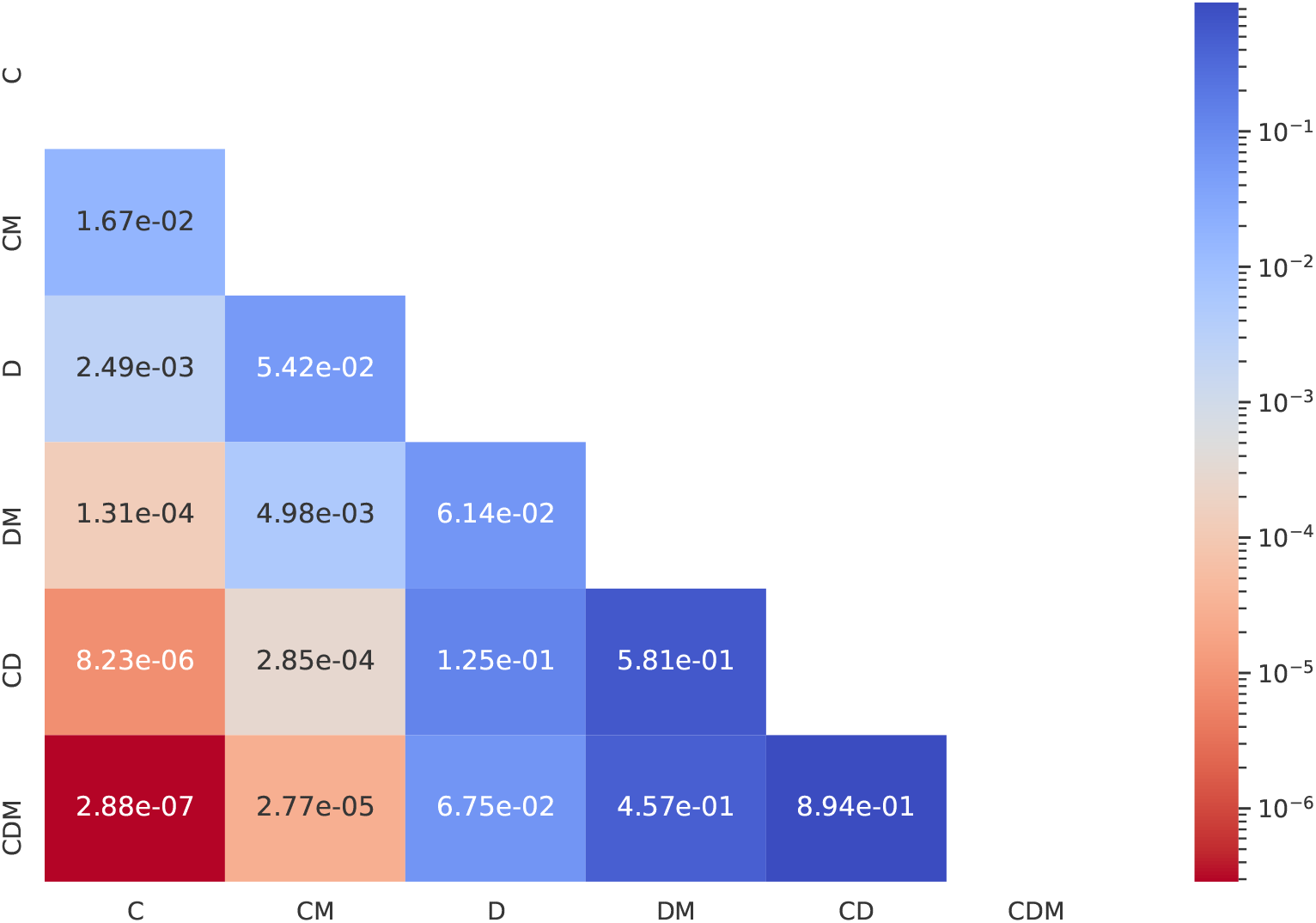
Evaluating the statistical significance of each input data modality’s contribution in improving the management decision prediction MGMT_pred, multi_. ‘C’, ‘D’, and ‘M’ refer to clinical image, dermoscopic image, and patient metadata respectively, and the row and the column names refer to the experiments in the ablation study presented in Table 4. For each pair of experiments (i) and (j), the cell (i, j) contains the *p*-value corresponding to the McNemar’s test performed on the corresponding pair of predictions.

1. **Dermoscopic images may be more useful than clinical images for predicting management decisions**. We compare the experiments where dermoscopic images and clinical images are used without (‘D’ versus ‘C’) and with (‘DM’ versus ‘CM’) the patient metadata. When not using the metadata (‘D’ versus ‘C’), using dermoscopic images over clinical images significantly improves all the metrics by 7.94 ± 1.87% (*p* = 2.49*e* − 03). Similarly, in the presence of metadata (‘DM’ versus ‘CM’), using dermoscopic images significantly improves all the metrics by 8.11 ± 2.36% (*p* = 4.98*e* − 03) as compared to when using clinical images.
2. **The value of adding a clinical image is questionable when a dermoscopic image is already present**. We compare the experiments where clinical images are added in addition to a dermoscopic image, both in the absence (‘CD’ versus ‘D’; *p* = 1.25*e* − 01) or presence (‘CDM’ versus ‘DM’; *p* = 4.57*e* − 01) of patient metadata and observe no consistent pattern of either improvement or degradation in the metrics.
3. **The inclusion of patient metadata may improve the management prediction accuracy**. When using only clinical images (‘CM’ versus ‘C’), only dermoscopic image (‘DM’ versus ‘D’), or both (‘CDM’ versus ‘CD’), all but one metrics improved with the inclusion of metadata by 2.23 ± 2.68%, with the most impactful contribution of metadata being in the 10.63% improvement of sensitivity in ‘CDM’ versus ‘CD’, and the only metric which decreased was the precision in ‘CDM’ versus ‘CD’ (2.59% decrease). However, these improvements are not statistically significant with *p* = 1.67*e* – 02 (‘CM’ versus ‘C’), *p* = 6.14*e* − 02 (‘DM’ versus ‘D’), and *p* = 8.94*e* − 01 (‘CDM’ versus ‘CD’).

While predicting management decisions, we posit that the clinical penalty of misclassifying certain management decisions is more severe than others. For example, consider a lesion where the correct management decision is for the lesion to be excised. Incorrectly predicting a management decision of ‘no further examination’ when the lesion should be excised is a more severe mistake than predicting a management decision of ‘clinical follow up’, since the decision to excise may be corrected in a future examination. We can extend this assumption to also include cases where the model predicts NONE when the target label is CLNC. For example, an EXC or a CLNC misclassified as a NONE is a more severe mistake than a NONE misclassified as an EXC or a CLNC, because in the latter scenario, the best course of action can ultimately be determined by the dermatologist in the clinical visit.

Since the multi-task model has also been trained to predict lesion diagnosis, the confusion matrix for the diagnosis prediction task is shown in Figure 3(d). Looking at the relationship between the diagnosis and the management labels (Table 1), we notice that all the malignant skin lesions, namely melanomas (MEL) and basal cell carcinomas (BCC), map to the same management label, i.e., excision (EXC). This means that if we can accurately predict a lesion to be either BCC or MEL, we can infer that it has to be excised. Therefore, if we were to first diagnose skin lesions and then infer their management, we would misclassify 46 malignant cases (the number of BCC or MEL misclassified as neither BCC nor MEL; Figure 3(c)), and thus incorrectly predict their management. On the other hand, if we directly predict the management decisions, we only misclassify 3 malignant cases (1 BCC and 2 MEL; Figure 3(a)).

### MClass-D Dataset

Next, we validate our trained prediction model on the publicly available MClass benchmark^30^. For this, we use the multi-task prediction model from Section “A multi-task prediction model” to simultaneously predict the diagnosis labels (DIAG) and the clinical management decisions (MGMT) for the 100 dermoscopic images in the MClass-D dataset. We use the multi-task model trained on the Interactive Atlas of Dermoscopy as is and do not fine-tune on the MClass-D dataset.

The prediction classes for DIAG are benign (BNGN) or malignant (MLGN), whereas those for MGMT are excision (EXC) or not (NOEXC). While the diagnosis ground truth labels from the ISIC Archive are available for the lesions, there are multiple ways of choosing a target label for the clinical management decision. Therefore, we look at two possible ways of assigning the “ground truth” management decision: using the aggregated recommendations of the 157 dermatologists present in the dataset (MGMT_GT, agg_), or using the the diagnosis ground truth to derive the “true” management decision (MGMT_GT, true_) (where “true” indicates the ideal management decision if the underlying diagnosis was known). For each of the two scenarios, we compare the performance of the directly predicted management decision (MGMT_pred_) to that of a scenario when the predicted diagnosis is used to infer the management decision (MGMT_infr_), similar to Section “Predicting whether a lesion should be excised or not”.

The confusion matrices and the ROC curves for these two sets of predictions (MGMT_infr_ and MGMT_pred_) as compared to both methods of choosing the “ground truth” management labels are presented in Figure 5(a) and (b) respectively. When we set MGMT_GT, agg_ as the target labels, as shown in the left column and red curves of Figure 5(a) and (b) respectively, we observe that predicting the management decision directly (MGMT_pred_) performs well for both the management classes without favoring any single particular class and achieves a notable improvement in the area under the ROC curve, as compared to when inferring the management decision (MGMT_infr_) based on the model’s diagnosis prediction. Additionally, as discussed in Section “A multi-task prediction model”, not all misclassification errors are equal, and the clinical penalty of misclassifying an EXC as NOEXC is much more than other errors. While an ROC curve shows the performance over all probability thresholds, the AUROC does not consider the actual decision of the model. When using a default probability threshold of 0.5, we note that directly predicting the management decisions incurs far fewer such mistakes than inferring the management (16 versus 36). Similarly, when setting MGMT_GT, true_ as the target management labels, we observe that although the area under the ROC curves are similar (Fig. 5(b) green curves), the confusion matrix (Fig. 5(a) right column) reveals that the MGMT_pred_ leads to better overall performance across both the classes and fewer instances of EXC being misclassified as NOEXC (6 versus 12).

**Figure 5.**
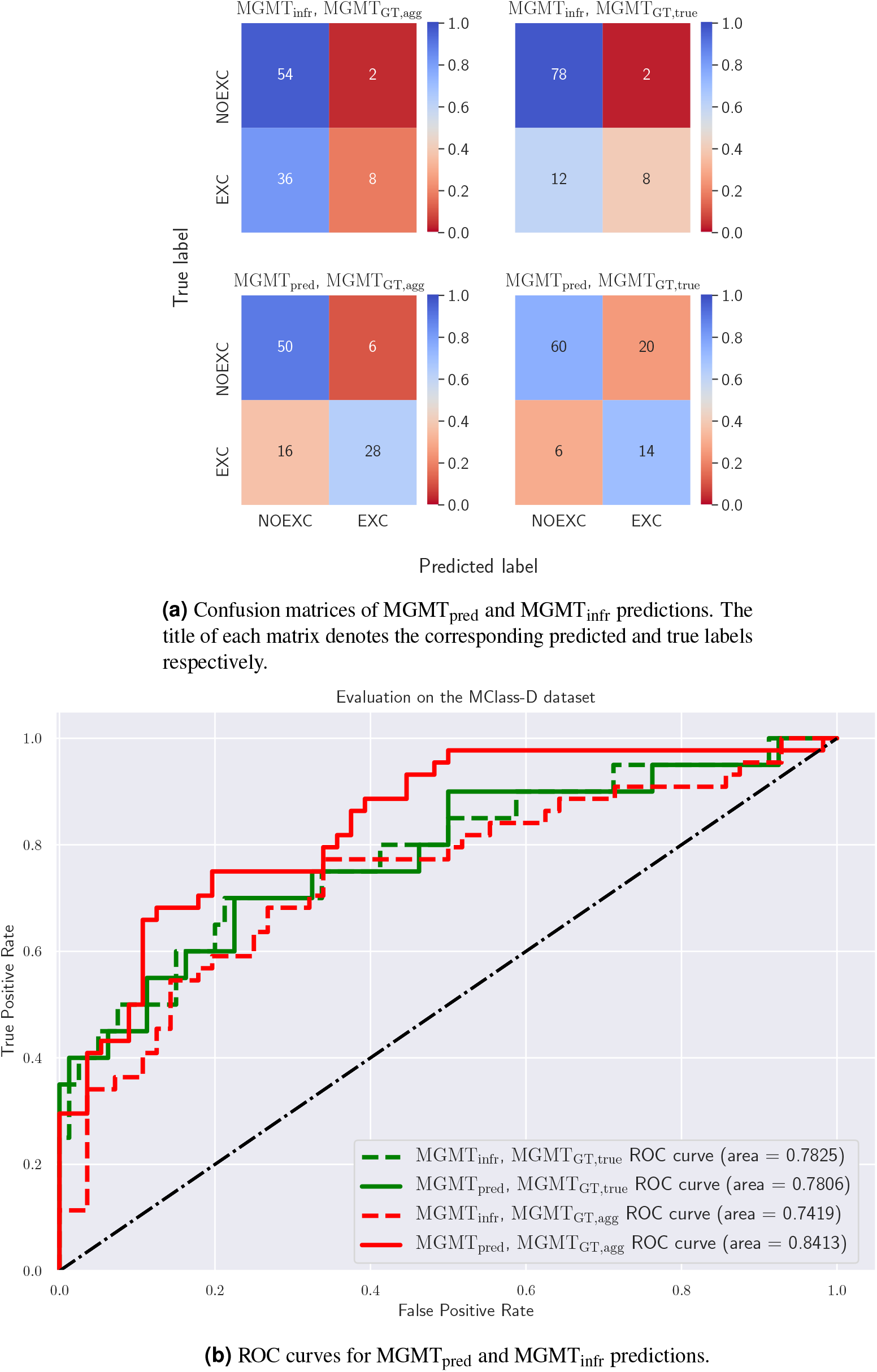
Evaluating the multi-task model on the MClass-D dataset. (a) Confusion matrices and (b) ROC curves for MGMT_pred_ and MGMT_infr_ predictions with both MGMT_GT, agg_ and MGMT_GT, true_ as target clinical management labels.

For evaluating the agreement between the model’s predictions and those of the 157 dermatologists, we calculate two agreement measures - Cohen’s kappa and Fleiss’ kappa. The Cohen’s kappa between our model’s predictions and that of the aggregated recommendations of the 157 dermatologists is 0.5424. This is higher than that of the agreement between all pairs of dermatologists (0.4124 ± 0.1032), and is comparable to the agreement between one dermatologist and the aggregated recommendations of all the others, repeated for all dermatologists (0.5497 ± 0.0899). Next, the Fleiss’ kappa for agreement among the recommendations of 157 dermatologists is 0.4086. To calculate the Fleiss’ kappa for capturing the agreement between our model’s predictions with those of the dermatologists, we calculate the agreement among a set of 156 dermatologists’ recommendations and the model’s predictions, and repeated by leaving out one dermatologist at a time, yielding a score of 0.4080 ± 0.0006. To address concerns that the recommendations of 156 dermatologists might overshadow the model’s predictions in the score calculated above, we repeat this experiment for a set of 10 predictions, comprising of 9 dermatologists’ recommendations and the model’s predictions, and repeat this 1000 times, yielding a score of 0.3961 ± 0.0301. These results indicate that our model’s clinical management predictions agree with those made by dermatologists as much as they do amongst each other.

Although Brinker et al.^18^ achieve a better performance at classifying melanomas than our model, we believe this can be attributed to multiple factors. First, Brinker et al. trained their prediction model on over 12,000 images and reported the mean of the results obtained from 10 trained models. Our model, on the other hand, is trained on considerably fewer images (413) and the reported results are from a single training run. Second, the training, validation, and testing partitions for Brinker et al. all come from the same data source, i.e., the ISIC Archive, whereas our model was trained on the Interactive Atlas of Dermoscopy and evaluated on images from the ISIC Archive, leading to a domain shift. CNNs have been shown to exhibit poor generalizability for skin lesion classification tasks when trained and evaluated on separate datasets^34^. Despite this, our multi-task prediction model is able to adapt to the new domain and exhibits strong generalization performance for clinical management predictions.

### Limitations

Although this study provides a proof of concept of the potential advantages of using deep learning to directly predict the clinical management decisions of skin lesions over inferring management decisions based on predicted diagnosis labels, it suffers from some limitations. First, the dataset that our model is trained on, the Interactive Atlas of Dermoscopy, only contains 20 diagnosis labels and 3 management labels and is not an exhaustive list of all diagnosis and management decisions. Second, although we trained the models on the Interactive Atlas of Dermoscopy with a reasonable effort on hyperparameter tuning and fine tuning, we did not pursue maximizing the classification accuracy. This means that even though our trained prediction model performs well on a held-out test set and is also able to generalize well when evaluated on data coming from a different source than the one it was trained on, better classification performance may be achievable with careful optimization of the prediction models. Finally, we acknowledge that unlike a dermatologist who has access to richer and non-image patient metadata such as patient history, demographics, patient preferences, and difficulty of diagnosis, our model only makes predictions based on the attributes present in these two datasets. However, this is not a technical limitation of our approach and rich multi-modal patient information can be incorporated as and when such attributes become available.

## Conclusion

In this work, we proposed a model to predict the management of skin lesions using clinical and dermoscopic lesion images and patient metadata. We showed that predicting the management decisions directly is significantly more accurate than predicting the diagnoses first and then inferring the management decision. Moreover, we also observed a considerable increase in the management prediction accuracy with a multi-task model trained to simultaneously predict the seven-point criteria, the diagnoses, and the corresponding management labels. Furthermore, evaluation of our model on another dataset showed excellent cross dataset generalizability and strong agreement with the recommendations of dermatologists.

Our goal with this work is not to propose a method that overrides the dermatologists, rather to provide a second opinion. Deep learning-based approaches for diagnosis, although commonplace as a clinical tool now^35–37^, were far from it a decade ago, and we predict a similar shift towards automated algorithms recommending the clinical management of diseases. Since we have proposed a learning-based approach, the model’s predictions can be made more robust and similar to dermatologists’ predictions by leveraging more complex patient attributes. Future research directions would include collecting and testing on other datasets with other skin conditions and treatments to assess the value of directly predicting management labels and deemphasizing the latent tasks such as diagnosis prediction.

## Materials and Methods

### Dataset

We have adopted the Interactive Atlas of Dermoscopy dataset^28^, a credible and extensively validated dataset that has been widely used to teach dermatology residents^38–40^, to train and evaluate our prediction models. The dataset contains clinical and dermoscopic images of skin lesions, patient metadata (patient gender and the location and the elevation of the lesion), the corresponding seven-point criteria^32^ for the dermoscopic images, and the diagnosis and the management labels for 1011 cases with mean [standard deviation] age of 28.08 [18.70] years; 489 males (48.37%); 294 malignant cases (29.08%); skin lesion diameter of 8.84 [5.39] mm. Following Kawahara et al.^28^, we split the dataset into training, validation, and testing partitions in the ratio of approximately 2 : 1 : 2 (413 : 203 : 395 to be precise) and maintain a similar distribution of the management labels across all the three subsets. A breakdown of the dataset according to the management and the diagnosis labels along with the details of the three splits is presented in Table 1, and more detailed breakdowns of the dataset according to the diagnosis classes and the patient metadata is presented as Supplementary Information (Supplementary Tables 1 and 2 respectively). We also present the evaluation of the multi-task prediction model on the MClass-D dataset^18^, a collection of 100 dermoscopic images from the ISIC Archive with the corresponding diagnosis labels and the clinical management decision of 157 dermatologists surveyed. The dermatologists came from 12 university hospitals in Germany and 43.9% of them were board-certified. The melanomas in the dataset were histopathology-verified and the nevi were diagnosed as benign either by expert consensus or by a biopsy.

### The prediction models

In this section, we present three management prediction models, a detailed breakdown of which is presented in Figure 6. In order to train prediction models that leverage both the clinical and the dermoscopic images as well as the patient metadata available in the dataset, we use a multi-modal framework^28^ and train two models: the first to predict the diagnosis and the second to predict the management decision. For both of these models, we adopt an InceptionV3^41^-backbone pretrained on the ImageNet dataset^42^ as the feature extraction model and drop the final output layer. We combine the extracted features from both clinical and dermoscopic images and compute the global average pooled responses, to which we then concatenate the patient metadata as a one-hot encoded vector. Next, we add a 1 × 1 convolutional layer for the prediction task (either the diagnosis or the management) as the final classification layer with the associated loss. We use the categorical cross-entropy loss to train the model, and they are denoted by *L*_DIAG_ and *L*_MGMT_ for the diagnosis and the management prediction models respectively. Since there is an inherent class imbalance in the dataset, we adopt a mini-batch sampling and weighting approach^28^. The loss function used to train these two single prediction task models is as follows:

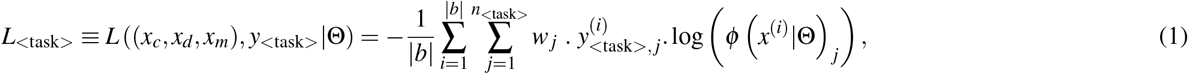

**Figure 6.**
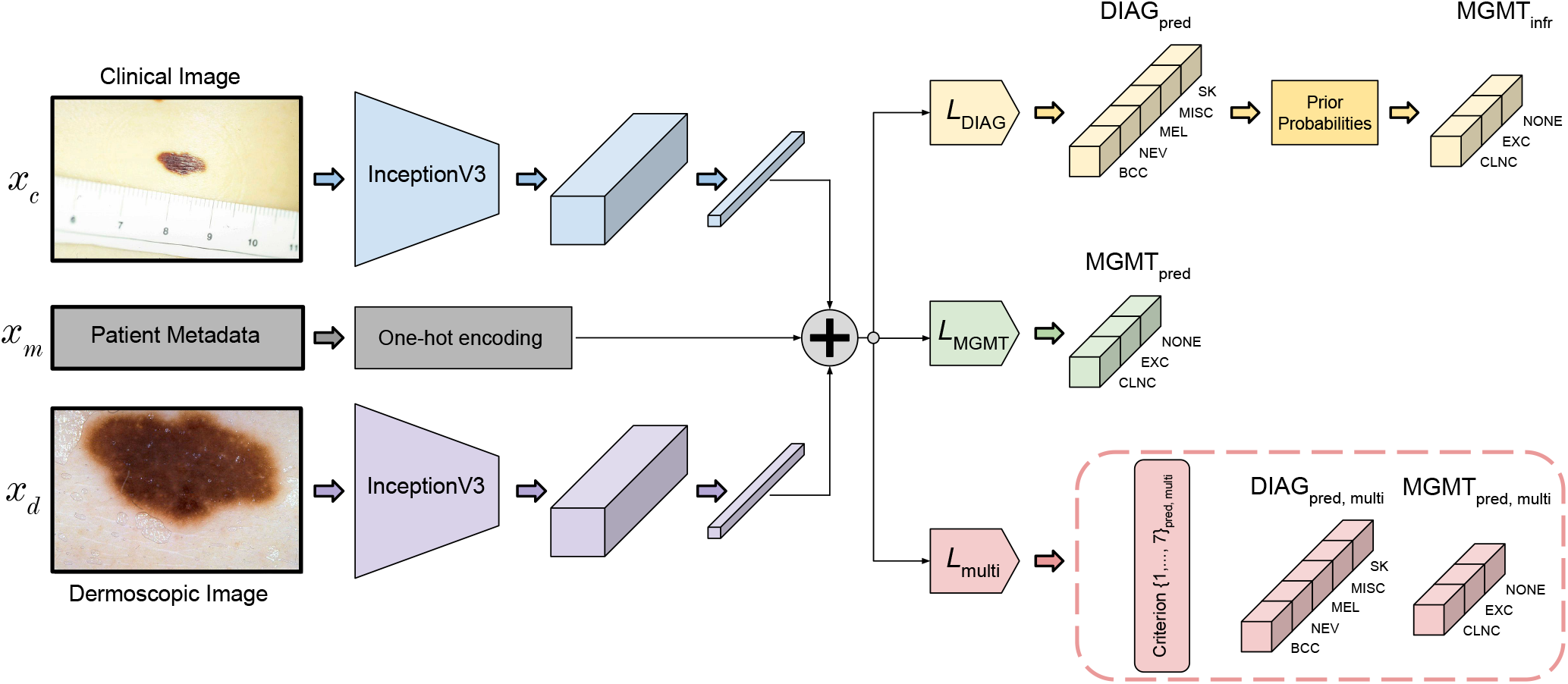
A breakdown of the inputs, outputs, loss functions, and architecture of the three prediction models. Global average pooled feature responses from the clinical and the dermoscopic images are extracted and concatenated (denoted by the plus symbol) with one-hot encoded patient meta-data, and the three models are trained with *L*_DIAG_, *L*_MGMT_, and *L*_multi_ respectively. The first model predicts the diagnosis labels (DIAG_pred_) which are then used along with the management priors to obtain inferred management decisions (MGMT_infr_), whereas the second model predicts the management decisions directly (MGMT_pred_). Finally, the last model is a multi-task one and is trained to predict the seven-point criteria, the diagnosis, and the management (outputs enclosed in the dashed box).

where *x*_*c*_, *x*_*d*_, *x*_*m*_ denote the clinical image, the dermoscopic image, and the patient metadata, respectively, |*b*| denotes the size of the mini-batch, ‘task’ denotes either the diagnosis or the management prediction task, and *y*_*<*task*>*_ and *n*_*<*task*>*_ denote the target variable and the number of classes for the corresponding tasks respectively. *w*_*j*_ denotes the weight assigned to the *j*^th^ class (calculated similar to Kawahara et al.^28^), and *ϕ*(*x*^(*i*)^|Θ) _*j*_ denotes the predicted probability for the *j*^th^ class given an input *x*^(*i*)^ by the model parameterized by Θ.

It has been shown that models optimized to jointly predict related tasks perform better on the individual tasks than models trained on each individual tasks separately^31,43^. Therefore, we train a third model by extending the multi-modal multi-task framework^28^ to simultaneously predict the seven-point criteria, the diagnosis, and the management decision. The architecture remains the same as the two models described above, except for the last layer, where we add a 1 × 1 convolutional layer for each prediction task as the final classification layer with the multi-task loss. The multi-task loss, denoted by *L*_multi_, accounts for all the 9 prediction tasks, namely: the seven-point criteria, the lesion diagnosis, and the lesion management, and is the sum of prediction losses for each of the tasks. As with the previous two models, we adopt the same mini-batch sampling and weighting approach. The loss function used to train this multi-task prediction model is defined as:

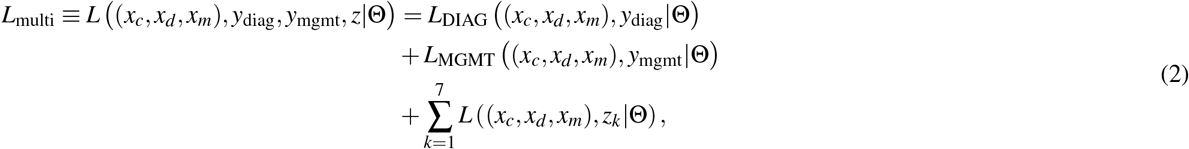

where *L*(·) denotes the categorical cross entropy loss (as described in Equation (1)) and Θ denotes the parameters of the multi-task model. The model outputs are *y*_DIAG_, *y*_MGMT_, and *z*_*k*_ ∈ ℤ7, which denote, respectively, the diagnosis label, the management label, and the integer score for each of the seven-point criteria.

### Making management predictions

#### Interactive Atlas of Dermoscopy Dataset

Since we ultimately seek the management decision for each patient, we evaluate all the models based on their management prediction performance. We examine two types of management decisions: predicting whether a lesion should be excised or not (our baseline) and predicting all the management decisions. The first model (Figure 1(a)) is trained to predict the diagnosis, and so we infer the management decisions MGMT_infr_ from its diagnosis predictions (DIAG_pred_) either by predicting the binary management decision MGMT_infr, binary_: EXC versus NOEXC (Figure 1 (a1)), or by predicting all management decisions MGMT_infr, all_, which for our dataset are EXC, CLNC, and NONE (Figure 1 (a2)). The second model is trained to predict the management decisions MGMT_pred_, either binary MGMT_pred, binary_ (Figure 1 (b1)) or all decisions MGMT_infr, all_, directly (Figure 1 (b2)). As for the third model, since it is trained to predict the diagnosis and the management along with the 7-point criteria (Figure 1(c)), we follow the same approach as the first two models to obtain management predictions. For all the prediction models, we also perform a three-fold cross validation to support the robustness of our results. The dataset was partitioned into three folds while ensuring that the class-wise proportions of the different categories (7-point criteria, diagnosis labels, and management decisions) remain similar across the training, validation, and testing partitions^28^. Moreover, in order to study the contribution of the three input data modalities (clinical image, dermoscopic image, and patient metadata) to the final management prediction, we also carry out an input ablation study on the multi-task prediction model (i.e., the third model; Figure 1(c)), where we train and evaluate six multi-task prediction models with different combinations of the three input modalities.

The binary management decisions, MGMT_infr, binary_ (Figure 1 (a1)) and MGMT_pred, binary_ (Figure 1 (b1)), are obtained using a binary mapping as described in Results and Discussion. Next, given that there are multiple ways to manage a disease category (e.g., in Table 1, NEV cases are managed using all three management labels), we adopt a data-driven approach (Figure 1 (a2)) to calculate the likelihood of all management decisions given a diagnosis prediction. We use the distribution of the management decisions across diagnosis classes in the training data to estimate the priors for assigning a management class *m*_*i*_ to a patient assigned the diagnosis class *d* _*j*_. This can be denoted as *p*(MGMT = *m*_*i*_|DIAG = *d* _*j*_). At inference time, given a patient’s data *x*, we estimate the probability of management *m*_*i*_ by marginalizing over all possible diagnosis classes:

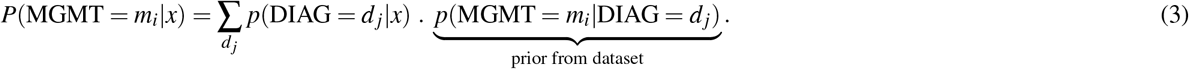

#### MClass-D Dataset

The multi-task model used to evaluate the images from MClass-D predicts both the lesion diagnosis (DIAG_pred_) and the clinical management (MGMT_pred_). The management labels inferred (MGMT_infr_) from the diagnosis predictions are obtained by the binary mapping described in Section “Predicting whether a lesion should be excised or not”. To recap, a lesion predicted to be malignant (MLGN) is mapped to the ‘excise’ (EXC) label and a lesion predicted to be benign (BNGN) would be mapped to ‘do not excise’ (NOEXC), meaning that the inferred management decision (MGMT_infr_) would have a direct mapping from the predicted diagnosis (DIAG_pred_).

Next, we look at the two different ways of obtaining the “ground truth” management labels. First, we aggregate the recommendations of the 157 dermatologists by majority voting to obtain a single prediction for each image (MGMT_GT, agg_), and use these as one type of target labels to compare the directly predicted management decisions (MGMT_pred_) and the inferred management decisions (MGMT_infr_). The second type of target labels are formed by generating the “true” clinical management labels by using a direct mapping from the disease diagnosis to clinical management. This is supported by the fact in an ideal world, we would want all malignancies (MLGN) to be excised (EXC) and all the benign lesions (BNGN) to not be (NOEXC). As such, the “true” clinical management labels (MGMT_GT, true_) are obtained by directly mapping the ground truth diagnosis classes to the corresponding management labels.

### Evaluation

For comparing the performance of the baseline binary labeling approach, we compare the per-class sensitivity averaged over the two classes for the two sets of binary predictions, MGMT_infr, binary_ and MGMT_pred, binary_.

Next, for each of the two sets of management predictions (MGMT_pred, all_ and MGMT_infr, all_), we obtain probabilistic predictions. To compare the performance of the two models, we choose to evaluate using two methods: (a) using the probabilistic management predictions, and (b) using the most likely label (i.e., choosing the single label with the highest predicted probability). While the evaluation for the latter is rather straightforward with accuracy values and confusion matrices, we formulate the following methodology for evaluating the quality of the probabilistic management predictions: given a set of predicted probability values (over management classes) and the corresponding target management labels, we report distance measures between the probabilistic predictions and the one-hot encoded representations of the target management labels.

### Statistical analysis

The primary outcome measures are class-wise sensitivity, specificity, precision, AUROC and overall accuracy for the diagnosis and the management prediction tasks.

To compare the probabilistic predictions for the management decision obtained using MGMT_infr, all_ and MGMT_pred, all_, we use four distance measures to compare the similarity of these probability-vectors to the one-hot-encoded target labels: cosine similarity, Jensen-Shannon divergence, Wasserstein distance, and Hellinger distance. Since a lower value is better for all these metrics except the cosine similarity, we instead use the (1 - cosine similarity) value for consistency across measures and visualize them using a swarm plot overlaid onto a box plot.

We use the two-sided Wilcoxon signed-rank test^44^ to compare the two sets of distance measures for each of the four measures since the differences between the two sets cannot be assumed to be normally distributed. We perform bootstrapping^45^ and sub-sampling 1000 times^46^ with a sample size of N/2 (where N is the size of the test set) with convergence criteria satisfied^47^. For all the distance measures, we report the means and the 95% confidence intervals along with Cohen’s d values^48^. Results are considered statistically significant at *p <* 0.001 level. For the ablation study, we use the mid-*p* McNemar’s test^49–51^ to compare the management prediction accuracies of the six models, where each model is trained with a different combination of input data modalities, and the results are considered statistically significant at *p <* 0.05 level.

For evaluation on the MClass-D dataset, we use two inter-rater measures for assessing the similarity of our model’s predictions with those of the 157 dermatologists: Cohen’s kappa^52^ and Fleiss’ kappa^53^. For Cohen’s kappa, we calculate the agreement between the model’s prediction and the labels obtained by aggregating the recommendations of all 157 dermatologists (MGMT_GT, agg_), and compare it with the average agreement between any two dermatologists. To account for the variability among the predictions of multiple dermatologists and how this might not be reflected in the aggregated recommendation, we also compare this with the agreement between one dermatologist and the aggregated recommendation of all others, repeating this over all 157 dermatologists in a leave-one-out fashion and report the average agreement. Unlike Cohen’s kappa, Fleiss’ kappa can assess the agreement among more than two raters, and therefore we first calculate the agreement among all the 157 dermatologists. For calculating the agreement of the model’s predictions with those of the dermatologists, we first calculate the Fleiss’ kappa for a set of 157 predictions obtained from 156 dermatologists and our model, and repeat this 157 times in a leave-one-out fashion and report the average agreement. However, this could lead to concerns that the agreement among the 156 dermatologists might affect the kappa value, so we further carry out the same experiment but with a set of 10 management decisions obtained from the recommendations of 9 dermatologists sampled at random from the dataset and our model’s predictions. We repeat this 1000 times and report the average agreement.

All statistical analyses were performed in Python using NumPy^54^, SciPy^55^, statsmodels^56^, PyCM^57^, and scikit-learn^58^ libraries, and all visualizations were created in Python using matplotlib^59^ and seaborn^60^ libraries.

### Implementation Details

The Keras framework^61^ was used to implement all the deep learning models. We follow a similar training paradigm as Kawahara et al.^28^. For all the models, the ImageNet-pretrained weights are frozen at the beginning and the models are fine-tuned with a learning rate of 10^−3^ for 50 epochs, followed by iteratively ‘un-freezing’ one Inception block at a time (starting from the Inception block closest to the output all the way to the second Inception block) and fine-tuning for 25 epochs with a learning rate of 10^−3^. We use real-time data augmentation using rotations, horizontal and vertical flipping, zooming, and height and width shifts for these initial 275 epochs. Lastly, we turn off data augmentation and fine-tune for 25 epochs. We use stochastic gradient descent with a weight decay of 10^−6^ and a momentum of 0.9 to optimize the weights.

## Data Availability

Both the datasets used in this article have been publicly released.

https://derm.cs.sfu.ca/

https://skinclass.de/mclass/

## Acknowledgements

K.A. is funded by Natural Sciences and Engineering Research Council of Canada (NSERC) grant RGPIN 06795 through a research assistantship. The authors are thankful to Ben Cardoen of the Medical Image Analysis Lab for discussions on Figure 2 and statistical analyses. The authors are grateful to the NVIDIA Corporation for donating a Titan X GPU used in this research.

## Author Contributions

K.A. worked on writing the code, performing the formal analysis and the experiments, and preparing the figures, with support from J.K. and G.H. K.A. worked on writing the initial draft. G.H. supervised the project, with support from J.K. All authors contributed to the design and the evaluation of the algorithm. All authors contributed to writing, reviewing, and editing the manuscript. All authors read and approved the Article.

## Data availability statement

Both the datasets used in this article have been publicly released:

1. The Interactive Atlas of Dermoscopy dataset^28,29^ is available at https://derm.cs.sfu.ca/.
2. The MClass-D dataset^18,30^ is available at https://skinclass.de/mclass/.

## References

1. Friedman, R. J., Rigel, D. S. & Kopf, A. W. Early Detection of Malignant Melanoma: The Role of Physician Examination and Self-Examination of the Skin. CA: A Cancer J. for Clin. 35, 130–151, DOI: 10.3322/canjclin.35.3.130 (1985).

2. Henning, J. S. et al. The CASH (Color, Architecture, Symmetry, and Homogeneity) Algorithm for Dermoscopy. J. Am. Acad. Dermatol. 56, 45–52, DOI: 10.1016/j.jaad.2006.09.003 (2007).

3. Bakheet, S. An SVM Framework for Malignant Melanoma Detection Based on Optimized HOG Features. Computation 5, 4, DOI: 10.3390/computation5010004 (2017).

4. Grzesiak-Kopeć, K., Nowak, L. & Ogorzałek, M. Automatic Diagnosis of Melanoid Skin Lesions Using Machine Learning Methods. In Rutkowski, L.et al. (eds.) International Conference on Artificial Intelligence and Soft Computing, 577–585, DOI: 10.1007/978-3-319-19324-3_51 (Springer, Cham, Zakopane, Poland, 2015).

5. Jaworek-Korjakowska, J. Computer-Aided Diagnosis of Micro-Malignant Melanoma Lesions Applying Support Vector Machines. BioMed Res. Int. 2016, 1–8, DOI: 10.1155/2016/4381972 (2016).

6. Murugan, A., Nair, S. H. & Kumar, K. P. S. Detection of Skin Cancer Using SVM, Random Forest and kNN Classifiers. J. Med. Syst. 43, 269, DOI: 10.1007/s10916-019-1400-8 (2019).

7. Oliveira, R. B., Pereira, A. S. & Tavares, J. M. R. Skin Lesion Computational Diagnosis of Dermoscopic Images: Ensemble Models based on Input Feature Manipulation. Comput. Methods Programs Biomed. 149, 43–53, DOI: 10.1016/j.cmpb.2017.07.009 (2017).

8. R D, S. & A, S. Deep Learning Based Skin Lesion Segmentation and Classification of Melanoma Using Support Vector Machine (SVM). Asian Pac. J. Cancer Prev. 20, 1555–1561, DOI: 10.31557/APJCP.2019.20.5.1555 (2019).

9. Esteva, A. et al. Dermatologist-level Classification of Skin Cancer with Deep Neural Networks. Nature 542, 115–118, DOI: 10.1038/nature21056 (2017).

10. Haenssle, H. et al. Man Against Machine: Diagnostic Performance of a Deep Learning Convolutional Neural Network for Dermoscopic Melanoma Recognition in Comparison to 58 Dermatologists. Annals Oncol. 29, 1836–1842, DOI: 10.1093/annonc/mdy166 (2018).

11. ISIC. ISIC 2019 Skin Lesion Analysis Towards Melanoma Detection (2019).

12. Kaggle.com. SIIM-ISIC Melanoma Classification | Kaggle (2020).

13. Hussain, M. A., Amir-Khalili, A., Hamarneh, G. & Abugharbieh, R. Segmentation-Free Kidney Localization and Volume Estimation Using Aggregated Orthogonal Decision CNNs. In Descoteaux, M.et al. (eds.) Medical Image Computing and Computer Assisted Intervention -MICCAI 2017, 612–620, DOI: 10.1007/978-3-319-66179-7_70 (Springer, Cham, Quebec City, Canada, 2017).

14. Lee, H. et al. Machine Friendly Machine Learning: Interpretation of Computed Tomography Without Image Reconstruction. Sci. Reports 9, 15540, DOI: 10.1038/s41598-019-51779-5 (2019).

15. Taghanaki, S. A. et al. Segmentation-free Direct Tumor Volume and Metabolic Activity Estimation from PET Scans. Comput. Med. Imaging Graph. 63, 52–66, DOI: 10.1016/j.compmedimag.2017.12.004 (2018).

16. Xue, W. et al. Direct Estimation of Regional Wall Thicknesses via Residual Recurrent Neural Network. In Niethammer, M.et al. (eds.) International Conference on Information Processing in Medical Imaging, 505–516, DOI: 10.1007/978-3-319-59050-9_40 (Springer, Cham, Boone, USA, 2017).

17. Zhao, R. et al. Direct Cup-to-Disc Ratio Estimation for Glaucoma Screening via Semi-Supervised Learning. IEEE J. Biomed. Heal. Informatics 24, 1104–1113, DOI: 10.1109/JBHI.2019.2934477 (2020).

18. Brinker, T. J. et al. Deep Learning Outperformed 136 of 157 Dermatologists in a Head-to-head Dermoscopic Melanoma Image Classification Task. Eur. J. Cancer 113, 47–54, DOI: 10.1016/j.ejca.2019.04.001 (2019).

19. Fujisawa, Y. et al. Deep Learning Surpasses Dermatologists. Br. J. Dermatol. 180, e39–e39, DOI: 10.1111/bjd.17470 (2019).

20. Hekler, A. et al. Effects of Label Noise on Deep Learning-Based Skin Cancer Classification. Front. Medicine 7, DOI: 10.3389/fmed.2020.00177 (2020).

21. Han, S. S. et al. Augmented Intelligence Dermatology: Deep Neural Networks Empower Medical Professionals in Diagnosing Skin Cancer and Predicting Treatment Options for 134 Skin Disorders. J. Investig. Dermatol. DOI: 10.1016/j.jid.2020.01.019 (2020).

22. Kroemer, S. et al. Mobile Teledermatology for Skin Tumour Screening: Diagnostic Accuracy of Clinical and Dermoscopic Image Tele-evaluation Using Cellular Phones. Br. J. Dermatol. 164, 973–979, DOI: 10.1111/j.1365-2133.2011.10208.x (2011).

23. Monheit, G. et al. The Performance of MelaFind. Arch. Dermatol. 147, 188, DOI: 10.1001/archdermatol.2010.302 (2011).

24. Wells, R., Gutkowicz-Krusin, D., Veledar, E., Toledano, A. & Chen, S. C. Comparison of Diagnostic and Management Sensitivity to Melanoma Between Dermatologists and MelaFind: A Pilot Study. Arch. Dermatol. 148, 1083, DOI: 10.1001/archdermatol.2012.946 (2012).

25. Carrara, M. et al. Multispectral Imaging and Artificial Neural Network: Mimicking the Management Decision of the Clinician Facing Pigmented Skin Lesions. Phys. Medicine Biol. 52, 2599–2613, DOI: 10.1088/0031-9155/52/9/018 (2007).

26. Marchetti, M. A. et al. Results of the 2016 International Skin Imaging Collaboration International Symposium on Biomedical Imaging challenge: Comparison of the accuracy of computer algorithms to dermatologists for the diagnosis of melanoma from dermoscopic images. J. Am. Acad. Dermatol. 78, 270–277.e1, DOI: 10.1016/j.jaad.2017.08.016 (2018).

27. Argenziano, G. et al. Interactive Atlas of Dermoscopy: A Tutorial (Book and CD-ROM) (2000).

28. Kawahara, J., Daneshvar, S., Argenziano, G. & Hamarneh, G. Seven-Point Checklist and Skin Lesion Classification Using Multitask Multimodal Neural Nets. IEEE J. Biomed. Heal. Informatics 23, 538–546, DOI: 10.1109/JBHI.2018.2824327 (2019).

29. Kawahara, J., Daneshvar, S., Argenziano, G. & Hamarneh, G. 7-Point Criteria Evaluation Database (2019).

30. Brinker, T. J. et al. Comparing Artificial Intelligence Algorithms to 157 German Dermatologists: The Melanoma Classification Benchmark. Eur. J. Cancer 111, 30–37, DOI: 10.1016/j.ejca.2018.12.016 (2019).

31. Zamir, A. R. et al. Taskonomy: Disentangling Task Transfer Learning. In 2018 IEEE/CVF Conference on Computer Vision and Pattern Recognition, 3712–3722, DOI: 10.1109/CVPR.2018.00391 (IEEE, Salt Lake City, USA, 2018).

32. Argenziano, G. et al. Epiluminescence Microscopy for the Diagnosis of Doubtful Melanocytic Skin Lesions. Arch. Dermatol. 134, DOI: 10.1001/archderm.134.12.1563 (1998).

33. Davis, J. & Goadrich, M. The Relationship between Precision-Recall and ROC Curves. In Proceedings of the 23rd International Conference on Machine learning - ICML ‘06, 233–240, DOI: 10.1145/1143844.1143874 (ACM Press, Pittsburgh, USA, 2006).

34. Yoon, C., Hamarneh, G. & Garbi, R. Generalizable Feature Learning in the Presence of Data Bias and Domain Class Imbalance with Application to Skin Lesion Classification. In Medical Image Computing and Computer Assisted Intervention - MICCAI 2017, 365–373, DOI: 10.1007/978-3-030-32251-9_40 (Springer International Publishing, 2019).

35. Abràmoff, M. D., Lavin, P. T., Birch, M., Shah, N. & Folk, J. C. Pivotal Trial of an Autonomous AI-based Diagnostic System for Detection of Diabetic Retinopathy in Primary Care Offices. NPJ Digit. Medicine 1, 39, DOI: 10.1038/s41746-018-0040-6 (2018).

36. BusinessWire. Zebra Medical Vision Secures a Fourth FDA Clearance for AI for Medical Imaging (2019).

37. FDA. FDA Permits Marketing of Artificial Intelligence-based Device to Detect Certain Diabetes-related Eye Problems (2018).

38. Carli, P. et al. Pattern Analysis, Not Simplified Algorithms, is the Most Reliable Method for Teaching Dermoscopy for Melanoma Diagnosis to Residents in Dermatology. Br. J. Dermatol. 148, 981–984, DOI: 10.1046/j.1365-2133.2003.05023.x (2003).

39. Johr, R. H. Interactive CD of Dermascopy. Arch. Dermatol. 137, 831–832 (2001).

40. Lio, P. A. & Nghiem, P. Interactive Atlas of Dermoscopy. J. Am. Acad. Dermatol. 50, 807–808, DOI: 10.1016/j.jaad.2003.07.029 (2004).

41. Szegedy, C., Vanhoucke, V., Ioffe, S., Shlens, J. & Wojna, Z. Rethinking the Inception Architecture for Computer Vision. In 2016 IEEE Conference on Computer Vision and Pattern Recognition (CVPR), 2818–2826, DOI: 10.1109/CVPR.2016.308 (IEEE, Las Vegas, USA, 2016).

42. Deng, J. et al. ImageNet: A Large-scale Hierarchical Image Database. In 2009 IEEE Conference on Computer Vision and Pattern Recognition, 248–255, DOI: 10.1109/CVPR.2009.5206848 (IEEE, Miami, USA, 2009).

43. Vesal, S., Patil, S. M., Ravikumar, N. & Maier, A. K. A Multi-task Framework for Skin Lesion Detection and Segmentation. In OR 2.0 Context-Aware Operating Theaters, Computer Assisted Robotic Endoscopy, Clinical Image-Based Procedures, and Skin Image Analysis, 285–293, DOI: 10.1007/978-3-030-01201-4_31 (Springer International Publishing, 2018).

44. Wilcoxon, F. Individual Comparisons by Ranking Methods. Biom. Bull. 1, 80, DOI: 10.2307/3001968 (1945).

45. Efron, B. Bootstrap Methods: Another Look at the Jackknife. In Kotz, S. & Johnson, N. L. (eds.) Springer Series in Statistics (Perspectives in Statistics), 569–593, DOI: 10.1007/978-1-4612-4380-9_41 (Springer, New York, NY, 1992).

46. Pattengale, N. D., Alipour, M., Bininda-Emonds, O. R., Moret, B. M. & Stamatakis, A. How Many Bootstrap Replicates Are Necessary? J. Comput. Biol. 17, 337–354, DOI: 10.1089/cmb.2009.0179 (2010).

47. Athreya, K. B. Bootstrap of the Mean in the Infinite Variance Case. The Annals Stat. 15, 724–731, DOI: 10.1214/aos/1176350371 (1987).

48. Cohen, J. Statistical Power Analysis for the Behavioral Sciences (Academic Press, 1977).

49. McNemar, Q. Note on the Sampling Error of the Difference between Correlated Proportions or Percentages. Psychometrika 12, 153–157, DOI: 10.1007/bf02295996 (1947).

50. Lancaster, H. O. Significance Tests in Discrete Distributions. J. Am. Stat. Assoc. 56, 223–234, DOI: 10.1080/01621459.1961.10482105 (1961).

51. Fagerland, M. W., Lydersen, S. & Laake, P. The McNemar Test for Binary Matched-Pairs Data: Mid-p and Asymptotic are better than Exact Conditional. BMC Med. Res. Methodol. 13, DOI: 10.1186/1471-2288-13-91 (2013).

52. Cohen, J. A Coefficient of Agreement for Nominal Scales. Educ. Psychol. Meas. 20, 37–46, DOI: 10.1177/001316446002000104 (1960).

53. Fleiss, J. L. Measuring Nominal Scale Agreement Among Many Raters. Psychol. Bull. 76, 378–382, DOI: 10.1037/h0031619 (1971).

54. van der Walt, S., Colbert, S. C. & Varoquaux, G. The NumPy Array: A Structure for Efficient Numerical Computation. Comput. Sci. & Eng. 13, 22–30, DOI: 10.1109/MCSE.2011.37 (2011).

55. Virtanen, P. et al. SciPy 1.0: Fundamental Algorithms for Scientific Computing in Python. Nat. Methods 17, 261–272, DOI: 10.1038/s41592-019-0686-2 (2020).

56. Seabold, S. & Perktold, J. Statsmodels: Econometric and Statistical Modeling with Python. In Proceedings of the 9th Python in Science Conference, DOI: 10.25080/majora-92bf1922-011 (SciPy, 2010).

57. Haghighi, S., Jasemi, M., Hessabi, S. & Zolanvari, A. PyCM: Multiclass Confusion Matrix Library in Python. J. Open Source Softw. 3, 729, DOI: 10.21105/joss.00729 (2018).

58. Pedregosa, F. et al. Scikit-learn: Machine Learning in Python. J. Mach. Learn. Res. 12, 2825–2830 (2011).

59. Hunter, J. D. Matplotlib: A 2D Graphics Environment. Comput. Sci. & Eng. 9, 90–95, DOI: 10.1109/MCSE.2007.55 (2007).

60. Waskom, M. et al. mwaskom/seaborn: v0.8.1 (September 2017), DOI: 10.5281/zenodo.883859 (2017).

61. Chollet, F. et al. Keras. https://keras.io (2015).

