## Supplementary Information for "Predicting the Clinical Management of Skin Lesions using Deep Learning"

### Dataset breakdown

In Supplementary Table 1, we present a detailed breakdown of the seven-point criteria evaluation dataset along with the diagnosis-wise groupings used to train the models, and in Supplementary Table 2, we present a breakdown of the patient metadata, namely the sex of the patient, and the elevation and the location of the lesion.

| Diagnosis | Count | Abbreviation | Diagnosis Total | Management | | |
|---|---|---|---|---|---|---|
| | | | | CLNC | EXC | NONE |
| basal cell carcinoma | 42 | BCC | 42 | 0 | 42 | 0 |
| blue nevus | 28 | NEV | 575 | 268 | 278 | 29 |
| clark nevus | 399 | | | | | |
| combined nevus | 13 | | | | | |
| congenital nevus | 17 | | | | | |
| dermal nevus | 33 | | | | | |
| recurrent nevus | 6 | | | | | |
| reed or spitz nevus | 79 | | | | | |
| melanoma | 1 | MEL | 252 | 0 | 252 | 0 |
| melanoma (in situ) | 64 | | | | | |
| melanoma (less than 0.76 mm) | 102 | | | | | |
| melanoma (0.76 to 1.5 mm) | 53 | | | | | |
| melanoma (more than 1.5 mm) | 28 | | | | | |
| melanoma metastasis | 4 | | | | | |
| dermatofibroma | 20 | MISC | 97 | 24 | 23 | 50 |
| lentigo | 24 | | | | | |
| melanosis | 16 | | | | | |
| miscellaneous | 8 | | | | | |
| vascular lesion | 29 | | | | | |
| seborrheic keratosis | 45 | SK | 45 | 4 | 10 | 31 |
| **Management Total** | - | - | - | 296 | 605 | 110 |

**Supplementary Table 1.** Breakdown of the seven-point criteria evaluation dataset by management and diagnosis labels and the training-validation-testing splits used to train the models.

| Label | Count | Label | Count |
|---|---|---|---|
| *Sex* | | *Location* | |
| Female | 522 | Abdomen | 125 |
| Male | 489 | Acral | 62 |
| *Elevation* | | Back | 281 |
| Flat | 448 | Buttocks | 21 |
| Nodular | 123 | Chest | 100 |
| Palpable | 440 | Genital areas | 8 |
| | | Head-neck | 92 |
| | | Lower limbs | 200 |
| | | Upper limbs | 122 |

**Supplementary Table 2.** Breakdown of the patient metadata of the seven-point criteria evaluation dataset.
